# Development and Validation of an Artificial Intelligence-based Pipeline for Predicting Oral Epithelial Dysplasia Malignant Transformation

**DOI:** 10.1101/2024.11.13.24317264

**Authors:** Adam J Shephard, Hanya Mahmood, Shan E Ahmed Raza, Anna Luiza Damaceno Araujo, Alan Roger Santos-Silva, Marcio Ajudarte Lopes, Pablo Agustin Vargas, Kris D. McCombe, Stephanie G. Craig, Jacqueline James, Jill Brooks, Paul Nankivell, Hisham Mehanna, Syed Ali Khurram, Nasir M Rajpoot

## Abstract

Oral epithelial dysplasia (OED) is a potentially malignant histopathological diagnosis given to lesions of the oral cavity that are at risk of progression to malignancy. Manual grading of OED is subject to substantial variability and does not reliably predict prognosis, potentially resulting in sub-optimal treatment decisions. We developed a Transformer-based artificial intelligence (AI) pipeline for the prediction of malignant transformation from whole-slide images (WSIs) of Haematoxylin and Eosin (H&E) stained OED tissue slides, named ODYN (*Oral Dysplasia Network*). ODYN can simultaneously classify OED and assign a predictive score (ODYN-score) to quantify the risk of malignant transformation. The model was trained on a large cohort using three different scanners (Sheffield, 358 OED WSIs, 105 control WSIs) and externally validated on cases from three independent centres (Birmingham and Belfast, UK, and Piracicaba, Brazil; 108 OED WSIs). Model testing yielded an F1-score of 0.96 for classification of dysplastic vs non-dysplastic slides, and an AUROC of 0.73 for malignancy prediction, gaining comparable results to clinical grading systems. With further large-scale prospective validation, ODYN promises to offer an objective and reliable solution for assessing OED cases, ultimately improving early detection and treatment of oral cancer.

## 1. Introduction

Oral epithelial dysplasia (OED) presents a significant challenge in the realm of oral pathology, where accurate diagnosis and early detection are paramount for effective intervention and prevention of malignant progression. OED is a potentially malignant histopathological diagnosis encompassing various lesions of the oral mucosa, typically manifesting as white (leukoplakia), red (erythroplakia) or mixed red-white (erythroleukoplakia) lesions^1,2^.

Histopathological grading of Haematoxylin and Eosin (H&E) stained tissue using the World Health Organisation (WHO, 2017^3^) classification system remains the current accepted practice for diagnosis and risk stratification of OED lesions. This is a three-tier system for grading OED into mild, moderate and severe grades based on the presence, severity and location of a wide range of cytological and architectural histological features (28 in total^4,5^). By its nature, this approach suffers from significant intra- and inter-observer variability and has poor predictive value for malignant transformation risk, potentially impacting on patient management. An alternate binary grading system, categorising lesions as low- or high-risk, based on the number of cytological and architectural features (as listed in the WHO criteria) aimed to improve the reproducibility of grading^6,7^. However, studies have shown significant variability and unreliability in grading using both systems, highlighting the need for a more objective and reproducible methods that can better predict malignant transformation risk in OED^8,9^.

To address challenges in subjectivity and misclassification of precancerous and cancerous lesions, there is a growing interest in leveraging advanced technologies, particularly deep learning (DL), which has seen extensive use in medical image analysis over the past decade^10–12^. Several state-of-the-art models, such as U-Net^13^ and DeepLab^14^, have been developed to perform image classification and segmentation. These models typically use convolutional neural networks (CNN), such as ResNet^15^, as feature extractors. Within digital pathology, many CNN-based algorithms have been developed for segmenting both tissue type and nuclear instances^16–19^. Further, weakly supervised methods have became popular choices for the analysis of histology images, enabling slide-level classification based on patch-level predictions. These methods typically divide WSIs into smaller patches, before using CNNs to extract patch-level features^20–22^. However, despite their success, CNN-based models have limitations such as high computational overhead and difficulty in capturing long-range dependencies in images, when being used for either segmentation or classification.

Transformers have gained widespread attention in recent years as they have been successfully applied in several natural language processing and computer vision tasks such as classification^23–25^. A typical Transformer encoder consists of a multi-head self-attention (MSA) layer, a multi-layer perceptron (MLP), and a layer normalisation (LN). The MSA layer empowers Transformers to capture long-range dependencies, making them a strong candidate for semantic segmentation in medical images^26–28^. Transformers, therefore, have the potential to overcome some of the limitations of traditional CNNs. However, only a handful of methods have applied Transformers for semantic segmentation in medical images^26,29^. Their application in histology has primarily been constrained to classification tasks^30,31^, with semantic segmentation left relatively unexplored. This raises the question of whether Transformers can be harnessed for semantic segmentation of histological images.

In this study, we aimed to develop a novel, weakly supervised, DL pipeline that could reliably and objectively segment and classify OED, whilst predicting the risk of malignant transformation in OED, using WSIs of H&E-stained OED slides. Specifically, we achieve this using interpretable nuclear features from dysplastic regions on the WSI. Moreover, we conduct a rigorous evaluation of the performance of our pipeline by comparing it to other state-of-the-art methods. To demonstrate the robustness and generalisability of our approach, we have developed our model using a large cohort with extended validation on unseen datasets acquired from three national and international centres (Birmingham and Belfast, UK, and Piracicaba, Brazil). For reproducibility, we make our code publicly available: https://github.com/adamshephard/odyn_inference.

## 2. Results

In this retrospective multi-centric study, we propose an innovative weakly supervised method for predicting the progression of OED lesions to malignancy. We additionally aim to produce a model that classifies oral tissue slides as being dysplastic vs non-dysplastic. We achieved this by analysing H&E-stained WSIs obtained from oral tissue biopsies, using a CNN, a Transformer and a MLP, in what we have called our Oral DYsplasia Network, “ODYN” (see Figure 1).

**Figure 1.**
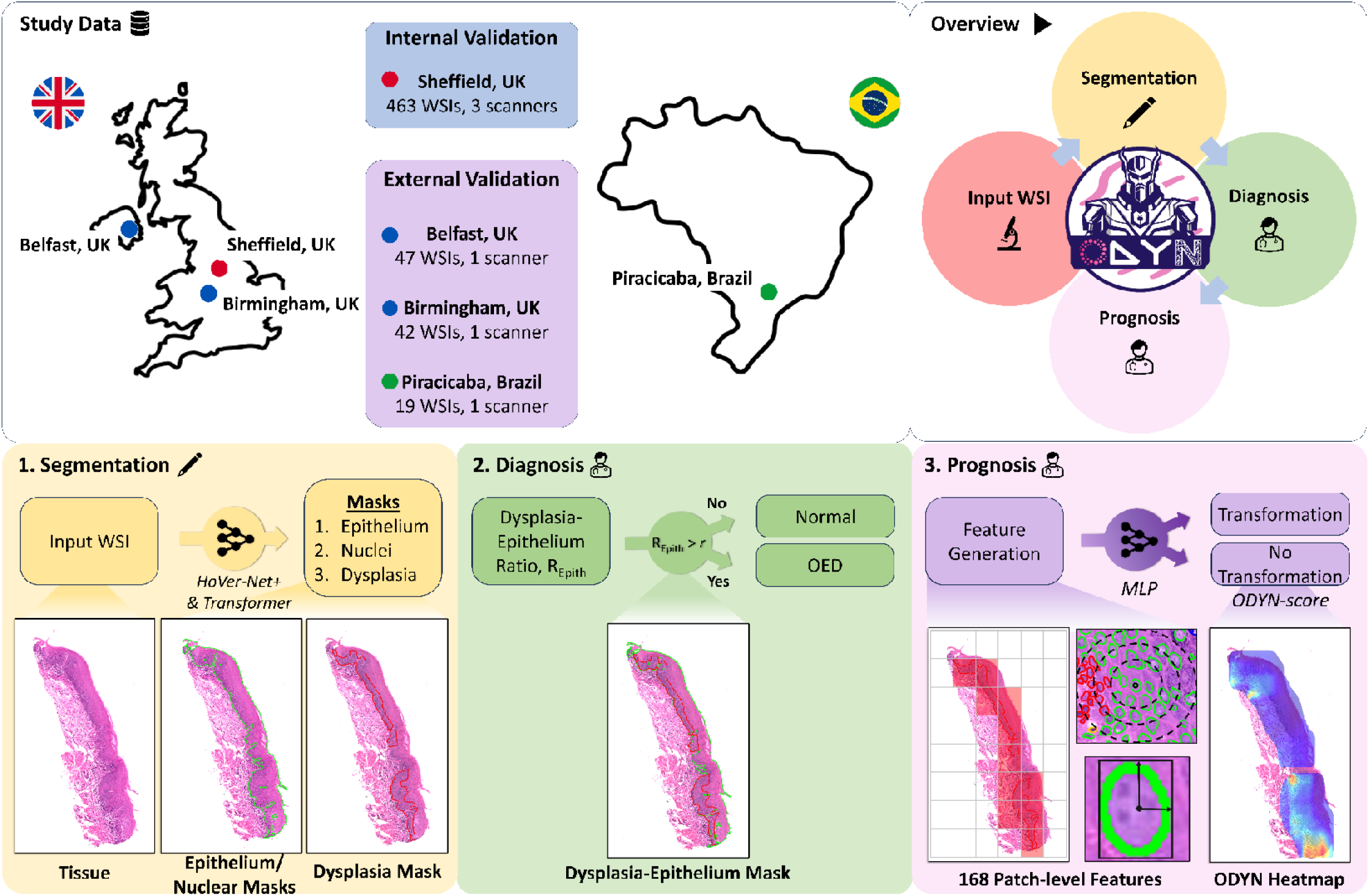
Overview of the ODYN pipeline. The top left panel shows the study data, whilst the top right panel shows an overview of the ODYN pipeline. The first stage (bottom left) takes an input oral tissue WSI and segments various tissue/cell types. This is done via HoVer-Net+ for epithelial and nuclei segmentation, and Trans-UNet to locate the dysplastic areas of the slide. The second step (bottom middle) diagnoses the input tissue as OED or normal, by calculating the ratio of the epithelium that is predicted to be dysplastic. If this is above a threshold (found on model training), then the slide is classified as OED. Finally, the third stage (bottom right) gives a prognosis, i.e. predicts whether the case will become cancerous. To do this, we generate patch-level nuclear features within the dysplastic regions alone and use these within a multi-layer perception (MLP), to predict malignant transformation.

### 2.1 Dysplasia Segmentation

In many cases of OED, histological atypia is not present across the entire tissue section, and thus, the first step of this work was to identify only the regions where dysplastic changes were present. We trained a Transformer (based on Trans-UNet^26^) to detect and segment the different dysplastic areas in each WSI. Internal testing of the ODYN dysplasia segmentation model demonstrated an F1-score of 0.81 (Recall = 0.85, Precision = 0.77) on OED cases and a specificity of 1.00 on controls. On external testing, the ODYN model achieved a F1-score of 0.71 (Recall = 0.76, Precision = 0.66). The results of the ODYN model were superior to that of other state-of-the-art methods including U-Net^13^, HoVer-Net+^16,32^, DeepLabV3+^33^, Efficient-UNet^34^, and Swin-UNet^29^ (see Supplementary Appendix, Table S2). Examples of dysplasia segmentation heatmaps are shown in Figure 2.

**Figure 2.**
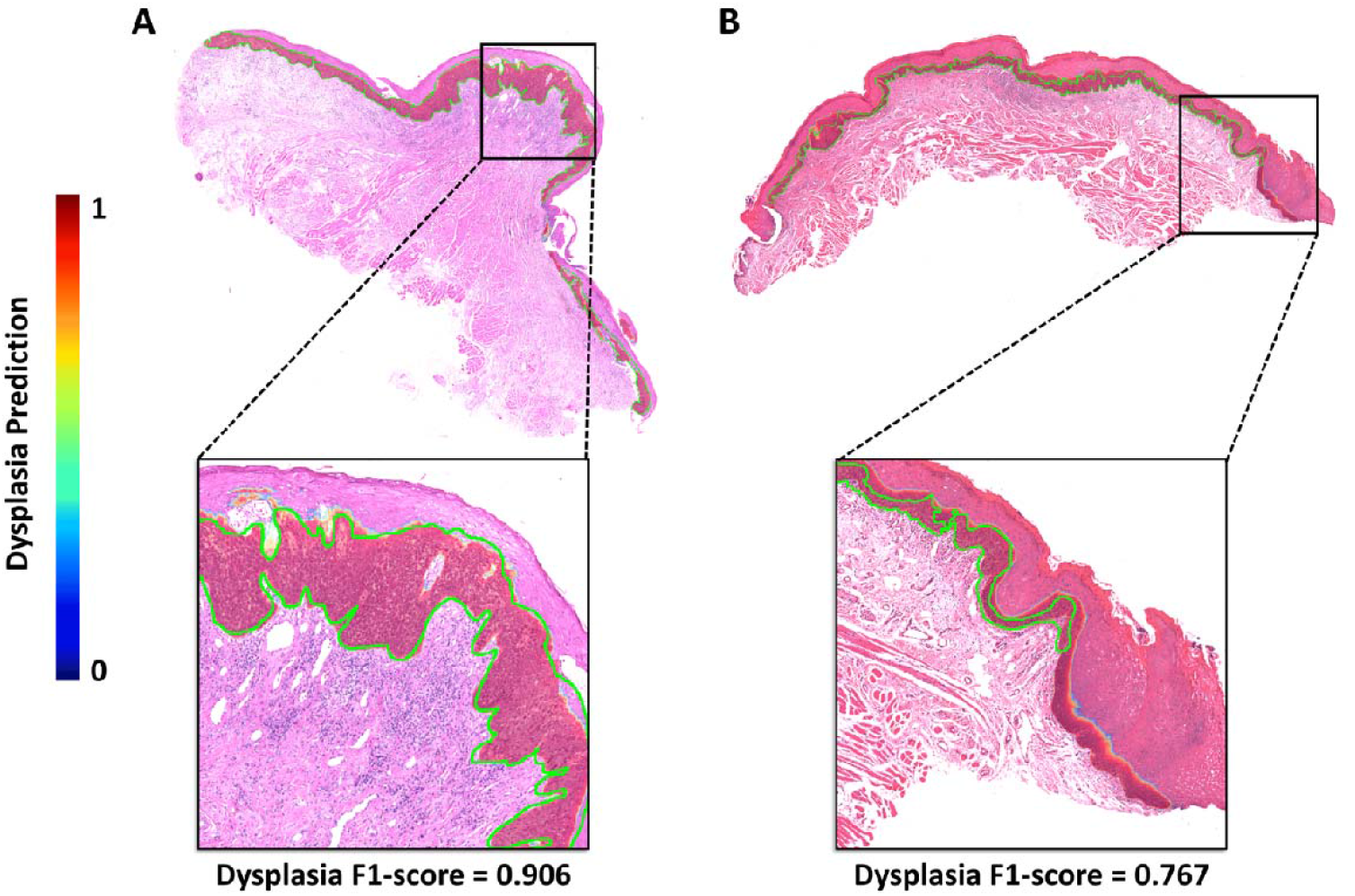
Dysplasia segmentation heatmap using the ODYN model. A) Severe OED (binary grade: high-risk) which transformed; B) Mild OED (binary grade: low risk) which did not transform. The green line depicts the ground truth dysplasia segmentation.

### 2.2 OED Classification

Next, we used a pretrained CNN (HoVer-Net+^16,32^) to segment the epithelium and the nuclei in the WSI. For OED classification, we calculate the proportion of the epithelium mask that was segmented as dysplastic (from the previous step) and use an empirically determined threshold to classify slides as being dysplastic vs. non-dysplastic. On internal testing, we achieved an F1-score of 0.96 (AUROC = 0.93, Recall = 0.94, Precision = 0.97). The performance remained high on external testing, gaining an F1-score = 0.96 (Recall = 0.93, Precision = 1.00), showing the robustness and generalisability of the proposed method.

### 2.3 Malignant Transformation Prediction

We generated patch-level morphological features in the dysplastic regions of OED cases, which were used as input to a MLP to calculate a risk-score for malignancy progression (the ODYN-score). On internal cross-validation we attained an AUROC of 0.71 for predicting malignant transformation, which remained relatively constant on external validation, rising to 0.73 (Table 1). These scores are competitive to existing clinical grading systems including WHO (2017) and binary grades. However, it must be noted that the binary grading system had a higher AUPRC of 0.72 when compared to the ODYN-score. For a complete evaluation, we also compared our ODYN-score to the other grading systems through a survival analysis (see Figure 3). On internal testing, our ODYN-score gained a comparable C-index of 0.65 and hazard ratio of 3.40, when compared to the other grading systems, and was shown to be significant (*p* < 0.001). On external testing the ODYN-score (C-index = 0.63) again attained comparable performance to both the binary grading system (C-index = 0.62), and WHO grading system (G2 stratification; C-index = 0.61), with all three being significant. The ODYN-score continues to surpass the WHO G1 stratification in terms of C-index and hazard ratio on both internal and external testing. Overall, these results show the prognostic significance and utility of the ODYN-score, being comparable to that of a pathologist’s binary grade for predicting transformation-free survival.

**Table 1.**
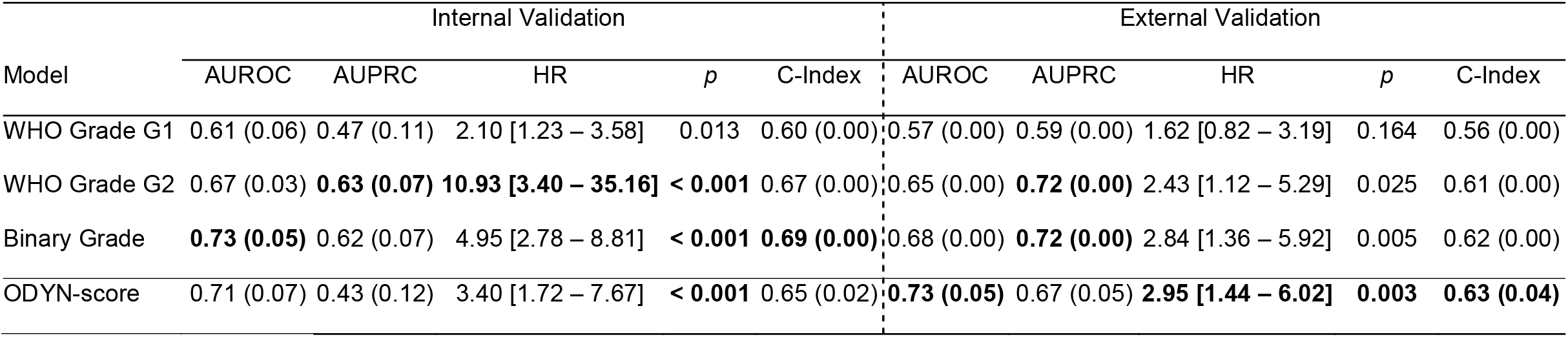
Slide-level results for transformation prediction. Here, WHO Grade G1 is mild/moderate vs severe cases, whilst WHO Grade G2 is mild vs moderate/severe cases. For AUROC, AUPRC and C-Index, the mean value is given with the standard deviation in brackets. For the hazard ratio, HR, we additionally provide the 95% confidence interval in square brackets.

**Figure 3.**
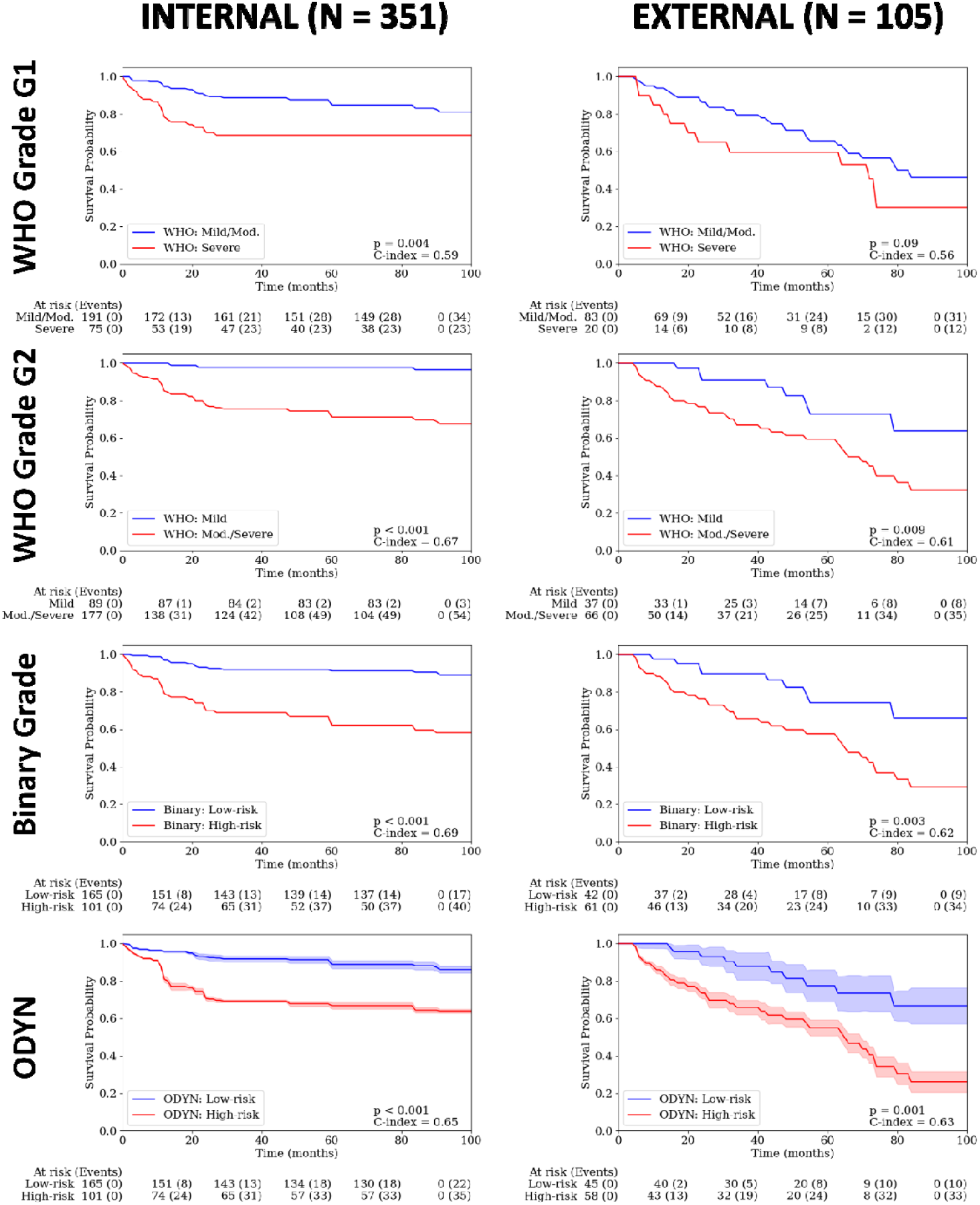
Kaplan-Meier transformation-free survival curves. Internal testing is on the left and external testing is on the right. The top row is WHO Grade G1 (i.e. Mild/Moderate vs Severe OED), second row is WHO Grade G2 (i.e. Mild vs Moderate/Severe OED), followed by the Binary grade and the ODYN-score.

### 2.4 Feature Analysis

For our feature analysis, we compared both nuclear counts and area ratio in the top ten patches from cases that ODYN predicted to transform (i.e., true positives, TPs) against those correctly predicted to not transform (i.e., true negatives, TNs). This analysis was performed on the external data alone, and boxplots are given in the Supplementary Appendix, Figure S2. The nuclear count analysis found a significantly higher number of “other” nuclei within the non-epithelial tissue (*d* = 1.31, *p* < 0.001), in TPs when compared to TNs. It also showed a significantly higher number of “other” nuclei within the epithelium (i.e. intra-epithelial lymphocytes, IELs) in TNs when compared to TPs, however with a small effect size (*d* = 0.31). It also displayed a significantly higher number of both dysplastic epithelial nuclei (*d* = 1.32, *p* < 0.001) and normal epithelial nuclei (*d* = 1.06, *p* < 0.001) within TNs when compared to TPs. The area ratio analysis found a significantly higher number of “other” tissue in TPs when compared to TNs (*d* = 1.26, *p* < 0.001). Finally, it also showed a significantly higher number of both dysplastic epithelium (*d* = 0.49, *p* < 0.001) and normal epithelium (*d* = 0.78, *p* < 0.001) within TNs compared to TPs.

## 3. Discussion

Several studies have explored the application of machine learning, including DL, to study OED. The general focus of these methods has been to segment the epithelium (and the nuclei), either manually or via DL models^16,32,35,36^. These segmentations have then been used in further DL models to predict grade or transformation^32,35,37^ or for pathologist-curated features based on digital images^38^. However, there has been little focus on segmenting dysplastic regions only for downstream prediction of malignant transformation.

In this study, we introduce ODYN, a novel Transformer-based pipeline for OED segmentation, classification and malignant transformation prediction. This pipeline has been developed using the largest and most diverse multicentric OED dataset to date, digitised using six different scanners. The results obtained through rigorous testing and validation demonstrate the effectiveness of our models in various aspects of OED analysis. The ODYN dysplasia segmentation performance has consistently outperformed other state-of-the-art DL models. We found only one other study to attempt dysplasia segmentation in OED^36^. The authors used a DeepLabV3+ model and evaluated it at the patch-level on moderate/severe cases from a single centre. Our study improved on this, using a new Transformer-based architecture evaluated at the WSI-level on all types of OED (mild, moderate and severe) from multiple centres, gaining higher F1-scores. Furthermore, the ODYN model has demonstrated good generalisability across external unseen datasets, indicating its robustness and applicability in diverse clinical settings. This highlights the potential of Transformer-based architectures in accurately delineating regions of dysplasia in H&E stained WSIs of oral epithelial tissue. This novel ground-breaking approach has the potential to redefine the landscape of OED diagnosis by providing more precise and consistent results.

ODYN has also demonstrated promising results for OED classification. In this study, we used the predicted dysplastic proportion of the epithelium in a WSI to determine a diagnosis of OED. We chose this method to classify a WSI as dysplastic, rather than classifying a WSI as dysplastic solely based on the presence of any predicted dysplasia. We made this choice because our model predictions often included small areas of false positives. This decision to define a threshold, proved to be successful on both internal and external testing. The high precision and recall achieved in classifying OED indicates the potential for automated diagnosis, which has the potential to increase diagnostic efficiency.

The application of ODYN-produced segmentation maps in predicting malignant transformation represents a significant advancement in computational pathology. Notably, this approach outperforms the *OMTscoring* pipeline proposed by Shephard *et al*. (2024)^32^ with a substantial improvement in AUROC score (see Supplementary Appendix, Table S3 for comparative results). However, some comments must be made regarding model performance on external testing. Despite the AUROC and AUPRC remaining high for ODYN, there was a substantial drop in C-index. This drop was also seen for the WHO and binary grades, suggesting that this may be attributed to differences between internal and external datasets (i.e. a domain shift). An analysis of the data used for external testing, showed a substantially different transformation-free survival rate for external centres. We see only 23% of cases to transform on internal testing. In contrast nearly, 42% of cases transformed in the external cohorts. This variation in the number of events is a clear indication of a type II prior (domain) shift between internal and external cohorts^39^ (see Supplementary Appendix, Figure S3, for Kaplan-Meier transformation-free survival curves), and is the clinical reality of retrospective cohorts.

The provided approach offers a significant level of explainability; a crucial aspect for translating computational models to clinical practice. Our model used morphological/spatial features within (and around) dysplastic areas to generate a prediction, thus emulating the features used by the pathologist in OED grading. Our feature analysis allowed the exploration of different nuclear types within dysplastic vs normal epithelium. These analyses showed, unsurprisingly perhaps, that more dysplastic nuclei were present in the patches that were predicted to transform (vs non transformed). Corroborating this, they additionally showed cases that were correctly predicted to not transform to have more normal epithelial tissue (and nuclei). Moreover, cases that transformed exhibited increased “other” nuclei in both connective tissue and the epithelium. We posit that this elevated density of “other” nuclei around the epithelium within transforming cases likely indicates the presence of peri-epithelial lymphocytes (PELs). Furthermore, emerging evidence from Bashir et al. (2023)^35^ highlights a higher density of PELs in cases undergoing malignant transformation. These findings align with previous research, noting increased immune cell infiltration in tongue lesions progressing to OSCC^40^ and identifying distinct immune-related subtypes in moderate and severe OED^41^.

We believe that the application of cutting-edge DL techniques, such as the ODYN pipeline, has huge translational potential which could help improve the accuracy and objectivity of OED diagnosis and grading. In addition to this, AI-based pipelines can improve prognostic reliability for prediction of cancer risk to improve patient outcomes. Future research should explore the scalability of the ODYN model to accommodate a broader range of oral conditions (such as those which can mimic OED) and tissue variations to assess whether it can accurately discriminate OED from other similar appearing conditions whilst still accurately predicting malignancy risk. This will enhance the clinical utility of the model and ultimately help provide more personalised patient care.

The authors acknowledge challenges and opportunities for future research based on this study. A potential challenge highlighted by this work is the need to address the interpretability of DL models in clinical practice. We have therefore used an interpretable model for transformation prediction, that considers known histological features (e.g., shape and size variations of nuclei) to generate predictions from dysplastic ROIs. We provide heatmaps for each slide to help explain model decisions. We believe such approaches can enhance trust and acceptance amongst healthcare professionals. The authors additionally acknowledge limitations related to a retrospective study. It would have been of interest to further explore the model performance for predicting OED recurrence. However, as there is no standardised treatment protocol for OED, there may have been variations in patient management between centres, and it is also difficult to reliably know the difference between true recurrence and field change. We would have additionally liked to incorporate social risk factors (e.g., smoking, alcohol consumption) in the multivariable modelling, however, it was not possible to acquire consistent information between the different centres. These issues could be addressed by a future prospective validation study. Despite this, the external validation of our models across multiple centres and scanners is a notable strength of this study. Future research could explore the application of ODYN in even more diverse clinical settings and expand its utility to other histopathological tasks beyond OED analysis. We suggest testing the method on other head and neck precancerous lesions, such as laryngeal dysplasia, as an interesting future direction of research.

In conclusion, our study signifies a substantial leap forward in the field of digital oral pathology, offering a powerful tool in ODYN for the detection, segmentation, and classification of OED, which we have made publicly available. This technology, underpinned by DL and Transformer-based architectures, showcases the potential of computational pathology to revolutionise the diagnosis and management of OED. The model’s exceptional performance in both internal and external testing, along with its ability to improve transformation prediction, underscores its potential to impact clinical practice positively. By addressing challenges and continuing to refine the model, we envision ODYN playing an important role in improving the diagnosis and management of OED and potentially other head and neck precancerous lesions in the future.

## 4. Methods

### 4.1 Study Cohorts

#### 4.1.1 Development and Internal Validation Cohort

The training cohort consists of a retrospective sample of histology tissue sections (dating 2008 to 2016 with minimum five-year follow-up data) collected from the Oral and Maxillofacial Pathology archive at the School of Clinical Dentistry, University of Sheffield, UK (referred to as the internal centre, hereafter). After microscopic inspection of the tissue sections by a Consultant Pathologist (SAK), newly cut 4 μm sections of the selected cases were obtained from formalin fixed paraffin embedded blocks and stained with H&E for analysis.

In total, 509 slides were collected from 406 patients. The slides were digitised to high-resolution WSIs at 40× objective power using one of three scanners: NanoZoomer S360 (Hamamatsu Photonics, Japan; 0.2258 mpp), Aperio CS2 (Leica Biosystems, Germany; 0.2520 mpp), Pannoramic 1000 (P1000, 3DHISTECH Ltd, Hungary; 0.2426 mpp). Further inspection of the WSIs excluded cases with poor staining quality, artefacts, distortions or blurring. The resulting cohort comprised 358 WSIs (n = 277 patients) with a confirmed histological diagnosis of OED and 105 WSIs (n = 81 patients) confirmed as non-dysplastic (controls). Due to incomplete follow-up data for five patients with OED (7 WSIs), these cases were only used for algorithm training and excluded from clinical outcome analysis. Thus, the final cohort included 351 WSIs (n = 272 patients) with confirmed diagnosis of OED amongst which 64 patients (79 WSIs) exhibited malignant transformation. Slides from the same subjects were assigned to the same fold during algorithm training/testing. An overview of the dataset and a CONSORT diagram are given in the Supplementary Appendix (Table S1 and Figure S1, respectively).

Clinical follow-up data for the OED cohort included patient age (at time of diagnosis), sex, intraoral site, OED grade (using binary and WHO 2017 systems) and transformation status. Transformation was defined as the progression of a dysplastic lesion to OSCC at the same clinical site within the follow-up period, and transformation time was measured in months. To ensure diagnostic consistency, all cases were evaluated by at least two certified pathologists (PMS, PMF, DJB, KH), who provided an initial diagnosis based on the WHO grading system (between 2008-2016). To confirm the WHO (2017) grade and assign binary grades, the cases were blindly re-evaluated by SAK and a clinician with a specialist interest and expertise in OED analysis (HM).

Amongst the 358 OED WSIs, HM exhaustively delineated regions of interest (ROI) representative of dysplasia in a large subset of 260 OED WSIs, using in-built annotation tools in the QuPath^®^ software^42^. Of the 105 control WSIs, HM additionally manually delineated the entire epithelium in a subset of 96 control WSIs^42^.

#### 4.1.2. Independent Validation Cohorts

The ODYN model was tested on three external datasets acquired from:

i. Precision Medicine Centre, Patrick G. Johnston Centre for Cancer Research, Queen’s University Belfast, UK (47 WSIs)
ii. Institute of Head and Neck Studies and Education, Institute of Cancer and Genomic Sciences, University of Birmingham, UK (42 WSIs)
iii. Oral Diagnosis Department, Semiology and Oral Pathology Areas, Piracicaba Dental School University of Campinas (UNICAMP), São Paulo, Brazil (19 WSIs)

Owing to the limited size of these datasets we combined them into a single multi-institutional external test set. Prior to the inclusion of external cases in the study, all WSIs were checked for suitability. Slides of poor quality, insufficient epithelium and cases positive for Candida Albicans or suggestive of Human Papilloma Virus infection were excluded. The WSI cohorts from Birmingham and Belfast were scanned at 40× objective power using a Pannoramic 250 (P250, 3DHISTECH Ltd., Hungary; 0.1394 mpp) and Aperio AT2 (Leica Biosystems, Germany; 0.2529 mpp) whole-slide scanner, respectively, to obtain digital WSIs. The Brazil cases were scanned at 20× objective power, by an Aperio CS (Leica Biosystems, Germany; 0.4928 mpp) scanner. The same clinical follow-up information was collected as that for the development/internal cohort. The external dataset did not include any control cases. Due to incomplete follow-up data for three patients with OED (3 WSIs), these cases were only used for algorithm validation and excluded from clinical outcome analysis. Thus, the final cohort included 105 WSIs (n=105 patients) amongst which 44 patients (44 WSIs) exhibited malignant transformation. A summary of this cohort and a CONSORT diagram are provided in the Supplementary Appendix (Table S1 and Figure S1, respectively). For model training, HM exhaustively delineated ROIs of dysplasia in 30 cases each from both Birmingham and Belfast, and an additional 18 cases from Brazil, using the QuPath^®^ software.

#### 4.1.3 Ethics Statement

Ethical approval for the study was obtained from the NHS Health Research Authority West Midlands (18/WM/0335), and experiments were conducted in compliance with the Declaration of Helsinki. Data collected were fully anonymised.

### 4.2 Analytical Workflow

#### 4.2.1 Dysplasia Segmentation

Since dysplastic changes may not be widespread across the entire tissue section in a slide, the first step of developing the DL pipeline involved identification and localisation of the dysplastic tissue regions for semantic segmentation. To achieve this, we trained a Transformer, based on Trans-UNet^26^, to automatically detect and segment the different dysplastic regions in each WSI across the training dataset. The model takes an input image of size 512 × 512 (at 1.0 micron per pixel, mpp, resolution) and outputs a dysplasia segmentation map.

For internal model testing, the dataset was split at 80/20, and controlled for both scanner type and OED grade. This resulted in 206 OED and 75 control WSIs in the training set, and 54 OED and 21 controls WSIs in the internal testing set, with ground truth annotations. Note, a higher proportion of controls were kept in the test set to ensure high specificity of OED segmentation in the control sample. After tessellating the WSIs and region masks into smaller patches (512 × 512 pixels, 184 pixels overlap, 10× magnification, 1.0 mpp), a total of 19,063 OED and 11,756 non-dysplastic patches were generated for model training/validation on the internal discovery cohort. This totalled 6,341 patches with ground truth annotations from the 78 WSIs in the external cohort. Various stain augmentation algorithms were tested during the development of the final model, using the TIAToolbox^43^. For the evaluation of OED segmentation, on both internal and external testing, large ROIs centred on the annotated tissue section were generated.

#### 4.2.2 OED Classification

A pretrained CNN-based HoVer-Net+^16,32^ model was used to segment the epithelium and the individual nuclei across each WSI. To classify OED, the proportion of the epithelium mask that was segmented as dysplastic was calculated and an empirically determined threshold used to classify slides as being dysplastic vs. non-dysplastic (dysplasia-epithelium ratio, R_Epith_). We found thresholds for this ratio based on all the WSIs used for training the dysplasia segmentation model (281 WSIs). We therefore tested the model internally on the remaining 182 WSIs, and externally on all 108 WSIs. HoVer-Net+ was used for inference alone for this task and was not further trained, as it is a state-of-the-art model for epithelium and nuclear segmentation and classification, that has been extensively pre-trained on OED data16,32.

#### 4.2.3 Malignant Transformation Prediction (ODYN-scoring)

The WSIs were tessellated into smaller patches (512 × 512 pixels, with 256 pixels overlap at 0.5 mpp) using tissue in the dysplastic regions alone. The nuclear segmentations from HoVer-Net+ were used to generate a total of 168 nuclear-based morphological and spatial features for each (dysplastic) patch. See the Supplementary Appendix, pp 7, for a list of the features used. These patch-level features were used as input to an MLP to calculate a risk-score for malignant transformation (ODYN-score). Thus, the ODYN-score indicated whether the algorithm predicted the case to have transformed (high-risk) or not transformed (low-risk). The MLP model had three layers with 168 nodes in the input layer, 64 nodes in the hidden layer, and 2 nodes in the output layer. It used a leaky ReLU activation function and dropout (0.2) after the hidden layer. The MLP was trained by Monte Carlo iterative-draw-and-rank sampling (IDaRS^20^), using a symmetric cross-entropy loss function and the Adam optimiser. This loss function was chosen as it has been shown previously to help overcome errors associated with weak labels^20,44^. IDaRS sampling was performed with parameter values of *k* = 5 for top predictive patches and *r* = 45 random patches, using a batch size of 256. On inference, the trained MLP calculated a prediction score for each patch in the dysplastic regions of the WSI, which can be considered the likelihood of a tile belonging to the positive class in the classification task (i.e., transformation). Slide-level scores were then obtained by taking the average prediction score across the top 50% ranked tiles. We used nuclear features with the aim of making the model interpretable. However, we additionally provided comparison to a ResNet34 classifier (trained with Macenko stain augmentation), using deep features, to show the impact on performance (see Supplementary Appendix, Table S3).

We used repeated five-fold cross-validation in our ODYN-scoring internal experiments based on the internal cohort. For each fold of cross-validation, we held one fold back for testing, and used the remaining four folds with a 90/10 split of data for training/validation. We then tested our model externally, by evaluating each model from internal cross-validation (i.e. all 15 folds) on the external data, and ensembling their predictions.

Survival analyses were additionally conducted to assess the prognostic significance of the ODYN-score in predicting transformation-free survival. Kaplan-Meier curves were generated, and log-rank tests were used to determine the statistical significance of grading (for ODYN-score, WHO and binary grades). A multivariate Cox proportional hazards model was employed, incorporating the ODYN-score, sex and age (and lesion site for internal testing), to predict transformation-free survival. We additionally performed this analysis using the binary and WHO grades in place of the ODYN-score for further comparison. Transformations were right censored at eight years across these analyses to ensure consistency between internal and external cohorts.

### 4.3 Feature Analysis

We generated nuclear counts and area ratios in the ten top-ranked tiles (as correctly predicted by iterative draw and rank sampling). For nuclear counts, we studied dysplastic epithelial nuclei, normal epithelial nuclei, “other” nuclei from within the epithelium (i.e., intra-epithelial lymphocytes, IELs), and “other” nuclei outside the epithelium (i.e., peri-epithelial lymphocytes, PELs). For area ratios, we studied the ratio of the patch that was “other” tissue, dysplastic epithelium, and normal epithelium. We performed paired two-tail *t*-tests (with false discovery rate, FDR correction for multiple comparisons) between patches from cases that ODYN correctly predicted to transform vs does not transform, to determine statistical significance. We additionally calculated effect sizes for these tests (cohen’s *d*).

### 4.4 Evaluation Metrics

Dysplasia segmentation performance (aggregated across all ROIs) was measured by calculating the F1-score, Recall and Precision. For internal testing of controls, a single measure of specificity for OED segmentation was reported, since a single incorrectly predicted pixel (e.g. incorrectly predicted as OED), would result in an F1-score, Recall, and Precision values of 0; thus, not giving an accurate representation of the model performance. For the evaluation of OED classification (dysplastic vs non-dysplastic) the F1-score, Recall, and Precision across all ROIs (and slides) were measured. Area under the receiving operating characteristic (AUROC) score was calculated for internal testing across all ROIs.

For the evaluation of the ODYN-scoring pipeline, we provide an AUROC score and an area under the precision-recall curve (AUPRC) score across all slides. We used concordance index (C-index) to measure the rank correlation between predicted risk scores and patients’ survival time. The reported C-index is the mean over each repeat of the experiment, whilst the p-value is calculated by two times the median p-value (from the log-rank test) over all repeats, to get a conservative estimate. We additionally used the hazard ratio (HR) and p-value output from the multivariate analyses as further metrics for evaluation. For reporting, we focus on the p-value from the multivariate analyses, being a more conservative and robust estimate. However, for completeness we also provide the log-rank p-value with the Kaplan-Meier curves.

## 5. Data Availability

We are unable to share the whole slide images and clinical data, due to restrictions in the ethics applications.

## 6. Code Availability

In the spirit of reproducibility, we have made the inference code for our pipeline available online, with model weights https://github.com/adamshephard/odyn_inference.

## 7. Acknowledgements

This study was supported by a Cancer Research UK Early Detection Project Grant, as part of the ANTICIPATE study (grant no. C63489/A29674) in addition to funding from the National Institute for Health Research (award no. NIHR300904). ALDA was funded by The São Paulo Research Foundation (grant no. 2021/14585-7). The authors express their gratitude to Professor Paul Speight (PMS), Professor Paula Farthing (PMF), Dr Daniel Brierley (DJB), and Professor Keith Hunter (KH) for their valuable contribution in providing the initial histological diagnosis. The authors would additionally like to thank Dr Mark Eastwood for his help with the visualisation of cases (and their segmentations) on the tiademos server (https://tiademos.dcs.warwick.ac.uk/).

## 8. Author Contributions

AS, Hanya M, SEAR, SAK and NMR designed the study with the help of all co-authors. AS, Hanya M and NMR developed the computational methods. AS wrote the code and carried out all the experiments. Hanya M and SAK provided the WSI annotations. SAK and Hanya M obtained the ethical approval and retrieved the histological and clinical data from Sheffield. KM, SC and JJ contributed to the collection of the histological and clinical data from Belfast. JB, PN and Hisham M contributed to the collection of the histological and clinical data from Birmingham. ALDA, ARSS, MAL, PAV contributed to the collection of the histological and clinical data from Brazil. All authors contributed to the writing of the manuscript.

## References

1. Speight, P. M., Khurram, S. A. & Kujan, O. Oral potentially malignant disorders: risk of progression to malignancy. Oral Surg. Oral Med. Oral Pathol. Oral Radiol. 125, 612–627 (2018).

2. Speight, P. M. Update on oral epithelial dysplasia and progression to cancer. Head Neck Pathol. 1, 61–66 (2007).

3. El-Naggar, A. K., Chan, J. K., Grandis, J. R. & Others. WHO classification of head and neck tumours. (2017).

4. WHO Classification of Tumours Editorial Board. Head and neck tumours [Internet; beta version ahead of print]. (International Agency for Research on Cancer, 2023).

5. Muller, S. & Tilakaratne, W. M. Update from the 5th Edition of the World Health Organization Classification of Head and Neck Tumors: Tumours of the Oral Cavity and Mobile Tongue. Head Neck Pathol. 16, 54–62 (2022).

6. Kujan, O. et al. Evaluation of a new binary system of grading oral epithelial dysplasia for prediction of malignant transformation. Oral Oncol. 42, 987–993 (2006).

7. Kujan, O. et al. Why oral histopathology suffers inter-observer variability on grading oral epithelial dysplasia: An attempt to understand the sources of variation. Oral Oncol. 43, 224–231 (2007).

8. Odell, E., Kujan, O., Warnakulasuriya, S. & Sloan, P. Oral epithelial dysplasia: Recognition, grading and clinical significance. Oral Dis. 27, 1947–1976 (2021).

9. Nankivell, P. et al. The binary oral dysplasia grading system: validity testing and suggested improvement. Oral Surg. Oral Med. Oral Pathol. Oral Radiol. 115, 87–94 (2013).

10. Litjens, G. et al. A survey on deep learning in medical image analysis. Med. Image Anal. 42, 60–88 (2017).

11. Madabhushi, A. & Lee, G. Image analysis and machine learning in digital pathology: Challenges and opportunities. Med. Image Anal. 33, 170–175 (2016).

12. Litjens, G. et al. Deep learning as a tool for increased accuracy and efficiency of histopathological diagnosis. Sci. Rep. 6, 1–11 (2016).

13. Ronneberger, O., Fischer, P. & Brox, T. U-net: Convolutional networks for biomedical image segmentation. in Lecture Notes in Computer Science (including subseries Lecture Notes in Artificial Intelligence and Lecture Notes in Bioinformatics) vol. 9351 234–241 (2015).

14. Chen, L.-C., Papandreou, G., Kokkinos, I., Murphy, K. & Yuille, A. L. Semantic Image Segmentation with Deep Convolutional Nets and Fully Connected CRFs. in International Conference on Learning Representations (ICLR) (2015). doi:10.1109/TPAMI.2017.2699184.

15. He, K., Zhang, X., Ren, S. & Sun, J. Identity mappings in deep residual networks. Lect. Notes Comput. Sci. (including Subser. Lect. Notes Artif. Intell. Lect. Notes Bioinformatics) 9908 LNCS, 630–645 (2016).

16. Shephard, A. J. et al. Simultaneous Nuclear Instance and Layer Segmentation in Oral Epithelial Dysplasia. Proc. IEEE/CVF Int. Conf. Comput. Vis. Work. October, 552–561 (2021).

17. Graham, S. et al. Hover-Net: Simultaneous segmentation and classification of nuclei in multi-tissue histology images. Med. Image Anal. 58, 101563 (2019).

18. Graham, S. et al. CoNIC Challenge: Pushing the frontiers of nuclear detection, segmentation, classification and counting. Med. Image Anal. 92, (2024).

19. Shephard, A. J. et al. An Automated Pipeline for Tumour-Infiltrating Lymphocyte Scoring in Breast Cancer. in 2024 IEEE International Symposium on Biomedical Imaging (ISBI) 1–5 (2024). doi:10.1109/ISBI56570.2024.10635302.

20. Bilal, M. et al. Development and validation of a weakly supervised deep learning framework to predict the status of molecular pathways and key mutations in colorectal cancer from routine histology images: a retrospective study. Lancet Digit. Heal. 3, e763–e772 (2021).

21. Kather, J. N. et al. Deep learning can predict microsatellite instability directly from histology in gastrointestinal cancer. Nat. Med. 25, 1054–1056 (2019).

22. Campanella, G. et al. Clinical-grade computational pathology using weakly supervised deep learning on whole slide images. Nat. Med. 25, 1301–1309 (2019).

23. Dosovitskiy, A. et al. An Image is Worth 16×16 Words: Transformers for Image Recognition at Scale. (2020).

24. He, K. et al. Transformers in medical image analysis. Intelligent Medicine vol. 3 59–78 (2023).

25. Vaswani, A. et al. Attention is all you need. in Advances in Neural Information Processing Systems vols 2017-Decem 5999–6009 (2017).

26. Chen, J. et al. TransUNet: Transformers Make Strong Encoders for Medical Image Segmentation. arXiv 1–13 (2021).

27. Dai, Y., Gao, Y. & Liu, F. TransMed: Transformers Advance Multi-Modal Medical Image Classification. Diagnostics 11, 1384 (2021).

28. Lin, A. et al. DS-TransUNet: Dual Swin Transformer U-Net for Medical Image Segmentation. IEEE Trans. Instrum. Meas. 71, 1–13 (2022).

29. Cao, H. et al. Swin-Unet: Unet-Like Pure Transformer for Medical Image Segmentation. Lect. Notes Comput. Sci. (including Subser. Lect. Notes Artif. Intell. Lect. Notes Bioinformatics) 13803 LNCS, 205–218 (2023).

30. Myronenko, A., Xu, Z., Yang, D., Roth, H. R. & Xu, D. Accounting for Dependencies in Deep Learning Based Multiple Instance Learning for Whole Slide Imaging. Lecture Notes in Computer Science (including subseries Lecture Notes in Artificial Intelligence and Lecture Notes in Bioinformatics) vol. 12908 LNCS (Springer International Publishing, 2021).

31. Vu, Q. D., Rajpoot, K., Raza, S. E. A. & Rajpoot, N. Handcrafted Histological Transformer (H2T): Unsupervised representation of whole slide images. Med. Image Anal. 85, 102743 (2023).

32. Shephard, A. J. et al. A fully automated and explainable algorithm for predicting malignant transformation in oral epithelial dysplasia. npj Precis. Oncol. 8, (2024).

33. Chen, L.-C., Papandreou, G., Schroff, F. & Adam, H. Rethinking Atrous Convolution for Semantic Image Segmentation. arXiv (2017).

34. Baheti, B., Innani, S., Gajre, S. & Talbar, S. Eff-UNet: A novel architecture for semantic segmentation in unstructured environment. IEEE Comput. Soc. Conf. Comput. Vis. Pattern Recognit. Work. 2020-June, 1473–1481 (2020).

35. Bashir, R. M. S. et al. A digital score of peri-epithelial lymphocytic activity predicts malignant transformation in oral epithelial dysplasia. J. Pathol. (2023) doi:10.1002/path.6094.

36. Liu, Y., Bilodeau, E., Pollack, B. & Batmanghelich, K. Automated detection of premalignant oral lesions on whole slide images using convolutional neural networks. Oral Oncol. 134, 106109 (2022).

37. Mahmood, H. et al. Development and validation of a multivariable model for prediction of malignant transformation and recurrence of oral epithelial dysplasia. Br. J. Cancer (2023) doi:10.1038/s41416-023-02438-0.

38. Mahmood, H. et al. Prediction of malignant transformation and recurrence of oral epithelial dysplasia using architectural and cytological feature specific prognostic models. Mod. Pathol. 35, 1151–1159 (2022).

39. Jahanifar, M. et al. Domain Generalization in Computational Pathology: Survey and Guidelines. arXiv 1–59 (2023).

40. Gannot, G., Gannot, I., Vered, H., Buchner, A. & Keisari, Y. Increase in immune cell infiltration with progression of oral epithelium from hyperkeratosis to dysplasia and carcinoma. Br. J. Cancer 86, 1444–1448 (2002).

41. Gan, C. P. et al. Transcriptional analysis highlights three distinct immune profiles of high-risk oral epithelial dysplasia. Front. Immunol. 13, 1–16 (2022).

42. Bankhead, P. et al. QuPath: Open source software for digital pathology image analysis. Sci. Rep. 7, 1–7 (2017).

43. Pocock, J. et al. TIAToolbox as an end-to-end library for advanced tissue image analytics. Commun. Med. 2, 120 (2022).

44. Wang, Y. et al. Symmetric cross entropy for robust learning with noisy labels. in Proceedings of the IEEE International Conference on Computer Vision vols 2019-Octob 322–330 (2019).

