## Supplementary Appendix for "Development and Validation of an Artificial Intelligence-based Pipeline for Predicting Oral Epithelial Dysplasia Malignant Transformation"

**Supplementary Tables**

Table S1. Overview of OED samples included in this study.

|  | Sheffield | Birmingham | Belfast | Brazil |
| --- | --- | --- | --- | --- |
| OED Cases, *n* | 277 | 47 | 42 | 19 |
| OED Slides, *n* | 358 | 47 | 42 | 19 |
| Median Age^a^ (IQR) | 63 (54 – 72) | 61 (50 – 70) | 62 (52 – 71) | 53 (45 – 58) |
| Sex, *n* (%) |  |  |  |  |
| Female | 167 (48) | 24 (51) | 21 (50) | 13 (68) |
| Male | 183 (52) | 23 (49) | 21 (50) | 6 (32) |
| Site, *n* (%) |  |  |  |  |
| Buccal Mucosa | 45 (13) | 6 (12) | 0 (0) | 7 (37) |
| Tongue | 158 (45) | 30 (64) | 29 (69) | 8 (42) |
| Floor of Mouth | 68 (19) | 3 (6) | 9 (21) | 0 (0) |
| Other | 80 (23) | 8 (17) | 4 (10) | 3 (16) |
| WHO grade, *n* (%) |  |  |  |  |
| Mild | 118 (33) | 24 (51) | 6 (14) | 11 (58) |
| Moderate | 134 (38) | 18 (38) | 25 (60) | 4 (21) |
| Severe | 99 (28) | 5 (11) | 11 (26) | 4 (21) |
| Binary grade, *n* (%) |  |  |  |  |
| Low-risk | 218 (62) | 28 (60) | 7 (17) | 11 (58) |
| High-risk | 133 (38) | 19 (40) | 25 (83) | 8 (42) |
| Transformation^b^, *n* (%) | 79 (23) | 10 (21) | 30 (71) | 4 (25) |
| Scanner, *n* (%) |  |  |  |  |
| Aperio CS2 | 180 (50) | 0 (0) | 0 (0) | 0 (0) |
| NanoZoomer S360 | 98 (27) | 0 (0) | 0 (0) | 0 (0) |
| P1000 | 80 (22) | 0 (0) | 0 (0) | 0 (0) |
| Aperio CS | 0 (0) | 0 (0) | 0 (0) | 19 (100) |
| P250 | 0 (0) | 47 (100) | 0 (0) | 0 (0) |
| Aperio AT2 | 0 (0) | 0 (0) | 42 (100) | 0 (0) |
| *Note.* *All provided statistics are at the slide-level.*  *^a^ Median age at OED diagnosis.*  *^b^ 5 Sheffield cases (7 WSIs) and 3 Brazil cases (3 WSIs) had no follow-up data and are excluded from the transformation figures.* | | | | |

Table S2. Comparative experiments for OED segmentation.

|  | Internal OED Cases  (n = 54) | | | Internal Controls  (n = 21) | External OED Cases  (n = 78) | | |
| --- | --- | --- | --- | --- | --- | --- | --- |
| Model | F1-score | Recall | Precision | Specificity | F1-score | Recall | Precision |
| U-Net^1^ | 0.775 | 0.796 | 0.755 | 0.996 | 0.685 | 0.694 | 0.676 |
| HoVer-Net+^2^ | 0.789 | 0.827 | 0.754 | 0.996 | 0.668 | 0.719 | 0.623 |
| DeepLabV3+^3^ | 0.802 | 0.817 | **0.788** | 0.998 | 0.704 | 0.704 | **0.705** |
| Efficient-UNet^4^ | 0.790 | 0.834 | 0.751 | 0.998 | 0.700 | **0.777** | 0.638 |
| Swin-UNet^5^ | 0.795 | 0.845 | 0.750 | 0.97 | 0.680 | 0.728 | 0.638 |
| **ODYN** | **0.807** | 0.845 | 0.773 | **0.998** | **0.708** | 0.764 | 0.660 |
| ODYN-SA | 0.805 | **0.858** | 0.758 | 0.997 | **0.708** | 0.744 | 0.676 |

Table S3. Slide-level results for transformation prediction. Here, WHO Grade G1 is mild/moderate vs severe cases, whilst WHO Grade G2 is mild vs moderate/severe cases. For AUROC, AUPRC and C-Index, the mean value is given with the standard deviation in brackets. For the hazard ratio, HR, we additionally provide the 95% confidence interval in square brackets.

|  | Internal Validation | | | | | External Validation | | | | |
| --- | --- | --- | --- | --- | --- | --- | --- | --- | --- | --- |
| Model | AUROC | AUPRC | HR | *p* | C-Index | AUROC | AUPRC | HR | *p* | C-Index |
| WHO Grade G1 | 0.61 (0.06) | 0.47 (0.11) | 2.10 [1.23 – 3.58] | 0.013 | 0.60 (0.00) | 0.57 (0.00) | 0.59 (0.00) | 1.62 [0.82 – 3.19] | 0.164 | 0.56 (0.00) |
| WHO Grade G2 | 0.67 (0.03) | **0.63 (0.07)** | **10.93 [3.40 – 35.16]** | **< 0.001** | 0.67 (0.00) | 0.65 (0.00) | **0.72 (0.00)** | 2.43 [1.12 – 5.29] | 0.025 | 0.61 (0.00) |
| Binary Grade | **0.73 (0.05)** | 0.62 (0.07) | 4.95 [2.78 – 8.81] | **< 0.001** | **0.69 (0.00)** | 0.68 (0.00) | **0.72 (0.00)** | 2.84 [1.36 – 5.92] | 0.005 | 0.62 (0.00) |
| ResNet34-IDaRS^6^ | 0.66 (0.08) | 0.34 (0.09) | 1.96 [1.08 – 3.67] | 0.042 | 0.59 (0.01) | 0.56 (0.07) | 0.48 (0.06) | 1.36 [0.66 – 2.80] | 0.400 | 0.52 (0.02) |
| OMTscore^7^ | 0.72 (0.08) | 0.46 (0.16) | 4.41 [2.08 – 9.78] | **< 0.001** | 0.68 (0.01) | 0.67 (0.05) | 0.58 (0.06) | 2.58 [1.19 – 5.58] | 0.016 | 0.60 (0.02) |
| **ODYN-score** | 0.71 (0.07) | 0.43 (0.12) | 3.40 [1.72 – 7.67] | **< 0.001** | 0.65 (0.02) | **0.73 (0.05)** | 0.67 (0.05) | **2.95 [1.44 – 6.02]** | **0.003** | **0.63 (0.04)** |

Table S4. Internal testing results with different loss functions and patch sizes/resolutions.

| Loss | Patch Size | Resolution (mpp) | OED cases | | | Controls |
| --- | --- | --- | --- | --- | --- | --- |
|  |  |  | F1-score | Recall | Precision | Specificity |
| Dice + CE | 356 | 1.0 | 0.794 | 0.824 | 0.767 | 0.998 |
| Dice + CE | 512 | 0.5 | 0.781 | 0.792 | 0.771 | 0.999 |
| **Dice + CE** | **512** | **1.0** | **0.807** | 0.844 | 0.773 | 0.997 |
| Dice | 512 | 1.0 | 0.795 | **0.852** | 0.746 | 0.996 |
| Jaccard | 512 | 1.0 | 0.000 | 0.000 | 0.000 | **1.000** |
| CE | 512 | 1.0 | 0.805 | 0.834 | **0.778** | 0.998 |
| Jaccard + CE | 512 | 1.0 | 0.784 | 0.828 | 0.744 | 0.996 |

Table S5. Internal testing on the OED cases and controls, whilst testing domain generalisation techniques.

| DG Method | OED cases | | | Controls |
| --- | --- | --- | --- | --- |
|  | F1-score | Recall | Precision | Specificity |
| WS | 0.798 | 0.839 | 0.760 | 0.998 |
| SA^8^ | 0.805 | **0.858** | 0.758 | 0.997 |
| DA^9^ | 0.682 | 0.723 | 0.644 | 0.984 |
| WS, SA | 0.802 | 0.851 | 0.758 | 0.997 |
| WS, DA | 0.700 | 0.749 | 0.657 | 0.991 |
| SA, DA | 0.735 | 0.774 | 0.701 | 0.992 |
| WS, SA, DA | 0.699 | 0.725 | 0.655 | 0.988 |
| Proposed | **0.807** | 0.845 | **0.773** | **0.998** |

**Supplementary Figures**

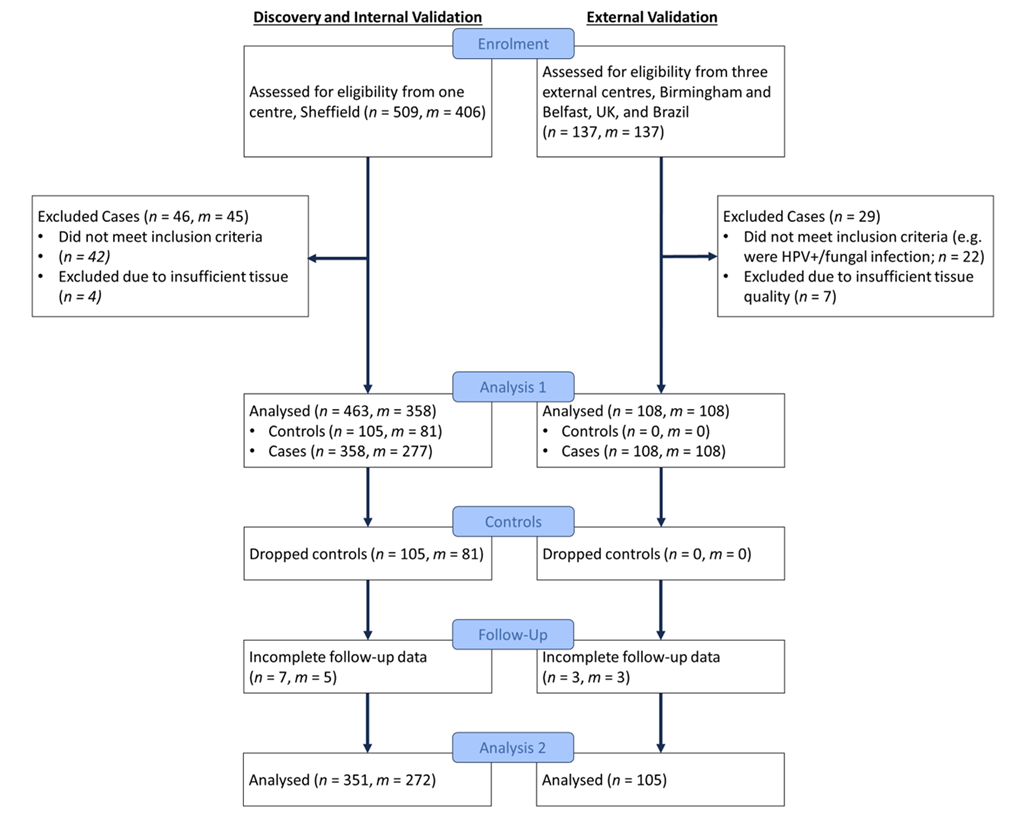

Figure S1. CONSORT flowchart illustrating samples. Here, n is the number of slides and m is the number of cases. Analysis 1 relates to the cases/controls available for training and evaluating ODYN for OED segmentation and classification. Analysis 2 relates to the cases available for training and evaluating ODYN for malignant transformation prediction. Since Analysis 2 requires follow-up data, 3 Brazil cases and 5 Sheffield cases were excluded from this analysis that were present in Analysis 1.

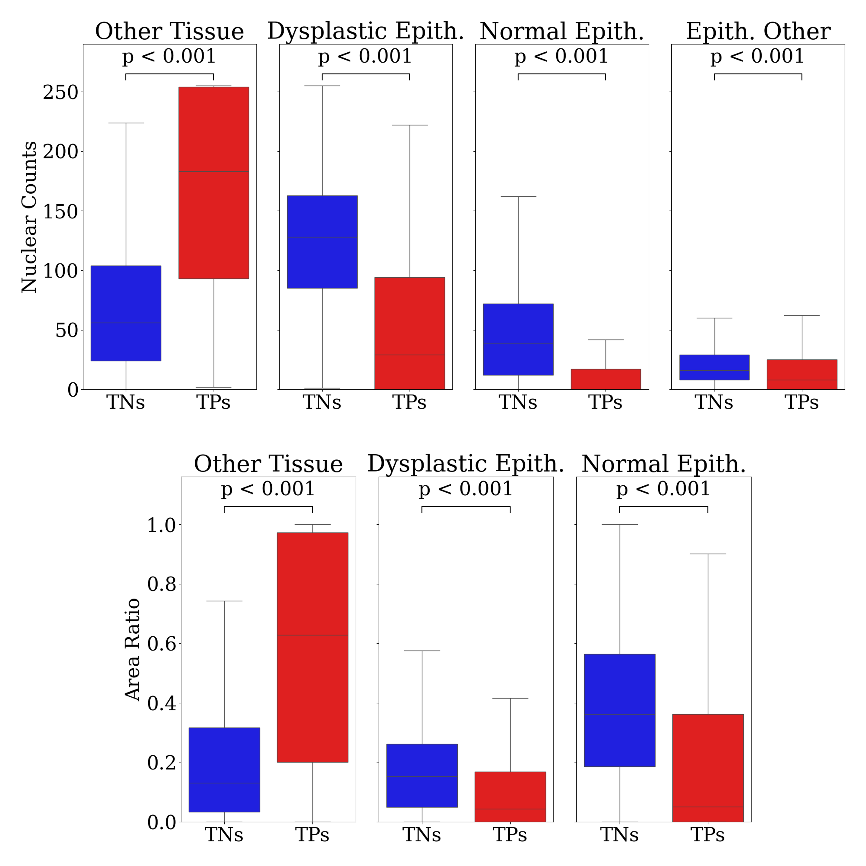

Figure S2. Boxplots showing the nuclear counts (top) and area ratios (bottom) for the top ten patches for true positives vs true negatives as predicted by ODYN for malignant transformation. For nuclear counts we have the number of “other” nuclei within the tissue that is not epithelium (left), the number of “other” nuclei within the epithelium (right), the number of epithelial nuclei within dysplastic epithelium (middle-left) and the number of epithelial nuclei within normal epithelium (middle-right). For area ratios we have the ratio of “other” tissue (left), the ratio of dysplastic epithelium tissue (middle) and the ratio of normal epithelium tissue (right) within a patch.

Figure S3. Kaplan-Meier transformation-free survival curves for the internal Sheffield and external datasets.

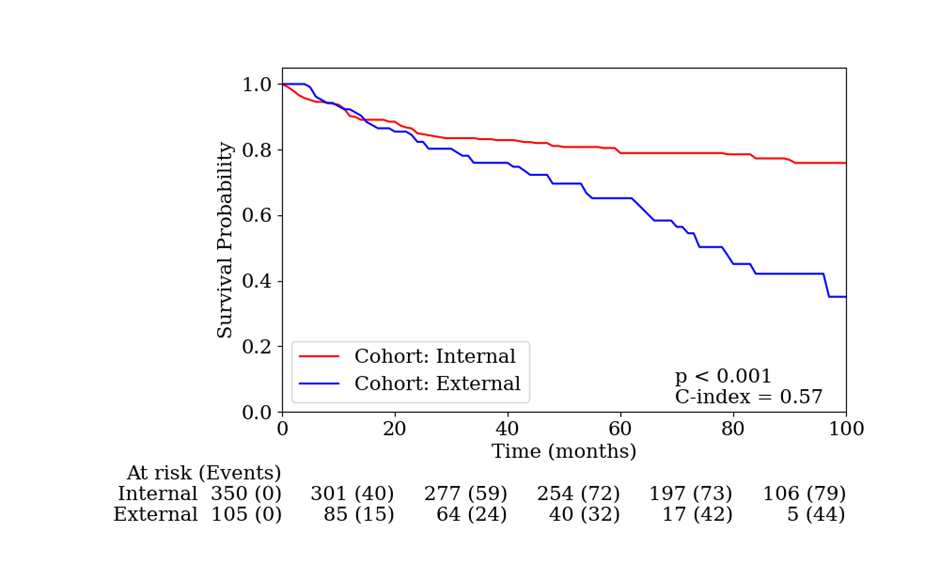

**Supplementary Methods**

***Dysplasia Segmentation***

We first tested the proposed model over varying patch sizes, resolutions, and loss functions. To aid our model in generalising to unseen domains, we also tested its performance based on various domain generalisation (DG) techniques. Methods we employed included: weighted sampling (WS), stain augmentation^8^ (SA), and domain adversarial training^9^ (DA). Following this, we compared our model against other state-of-the-art deep learning models for semantic segmentation, including Swin-UNet^5^, U-Net^1^ (ResNet-50^10^ backbone), Efficient-UNet^4^ (Efficient-Net-B7 backbone), DeepLabV3+^3^ (ResNet-101 backbone), and HoVer-Net+^11^ (segmentation decoder alone). All of these models were trained based on their default parameters, and pretrained on ImageNet.

We trained all models in two phases. We trained the decoders for 20 epochs first, before training the entire network for 30 epochs second. The Adam optimizer was used with a learning rate that decayed initially from 10^-4^ to 10^-5^ after 10 epochs, in both phases. We applied the following random data augmentations: flip, rotation, Gaussian blur, median blur, and colour perturbation. We additionally tested the effect of stain augmentation using the TIAToolbox^12^ implementation of the Macenko method^8^. This has been shown previously to help counter scanner-induced domain-shift^13–15^.

We found a patch size of 512×512 at 1.0 mpp, with a combined Dice and cross-entropy loss function to be best (see Table S4). To aid our model in generalising to unseen domains, we also tested its performance based on various domain generalisation (DG) techniques. These techniques yielded no improvement in performance on internal testing (see Table S5), with domain adversarial training hindering performance. Stain augmentation improved specificity on controls, with a slight reduction in F1-score. We suggest that these techniques were not beneficial on internal testing as slides from all three scanners were present in both the training and testing set. Instead, techniques such as stain augmentation may be more beneficial for external testing.

We compared our model to other state-of-the-art methods in Table S2. Here, we see the superiority of the proposed model (F1 = 0.81) when compared to all other models. DeepLabV3+ was the closest performing model (F1 = 0.80), with U-Net being worst (F1 = 0.78). The proposed model generalised well on external testing, gaining an F1-score of 0.71, and a high recall (see Table S2). Stain augmentation did not appear to improve the model F1-score; however, it did make the model more precise. We provide the comparative model results in Table S2, showing our proposed model to be best.

***Nuclear Features for Outcome Prediction***

After segmentation, each WSI was tessellated into smaller 512×512 tiles (20× magnification, 0.50 mpp) with 50% overlap. We used this tile size to ensure that each tile contained enough information for the prediction task, in line with previous studies^6,16^. We then generated tile-level features for use in a weakly supervised model for transformation prediction. For each tile, we calculated 104 morphological and 64 spatial features. The morphological features were obtained from 13 shape features for each nucleus in a tile (eccentricity, convex area, contour area, extent, perimeter, solidity, orientation, radius, major/minor axis, equivalent diameter, bounding box area/aspect ratio) with four tile-level statistics (mean, minimum, maximum, standard deviation) per nuclear type (epithelial and other). This resulted in 104 morphological features per tile. We computed the number of different nuclear types within a small radius of a nuclear instance, resulting in four counts per tile (number of epithelial nuclei around another nucleus, number of epithelial nuclei around epithelial nuclei, number of other nuclei around epithelial nuclei, and finally the number of other nuclei around other nuclei) over four varying radii (100, 200, 300 and 400 pixel radii). Finally, we took tile-level summary statistics (mean, minimum, maximum, standard deviation) across these 16 features, resulting in 64 spatial features per tile. We chose to use these 168 morphological/spatial features in preference to “deep” features output by CNNs, such as in traditional prediction tasks^6,17–19^, to offer transparency and explainability to the model inputs.

***Comparative Methods for Outcome Prediction***

We wished to further compare ODYN against other published benchmark weakly-supervised classification methods. Only two other methods have been proposed for malignant transformation prediction^7,16^. Bashir *et al.* (2023)^16^ used patches of tissue extracted from manually generated epithelial masks, and used these patches to train a ResNet-34 classifier for predicting malignant transformation. This model was trained using the iterative draw-and-rank sampling (IDaRS) method introduced by Bilal *et al.* (2021)^6^. Since, we produce dysplastic masks within this study using ODYN, we replicated this pipeline with our ODYN model, using this as a comparative study (here, labelled ResNet34-IDaRS). We employed stain augmentation during model training. Shephard *et al.* (2024)^7^ released the *OMTscoring* pipeline, which used HoVer-Net+^11^ to generate epithelial/nuclear segmentations. It then uses a multi-layer perceptron (MLP), based on nuclear features, to predict slide-level outcome (i.e. transformation). We have created ODYN to also replicate this pipeline and additionally produce the *OMTscore*. We therefore provide this as an additional comparative study. For the results of these comparative experiments, see Table S3.

**Supplementary References**

1.	Ronneberger, O., Fischer, P. & Brox, T. U-net: Convolutional networks for biomedical image segmentation. *Lect. Notes Comput. Sci. (including Subser. Lect. Notes Artif. Intell. Lect. Notes Bioinformatics)* **9351**, 234–241 (2015).

2.	Graham, S. *et al.* Hover-Net: Simultaneous segmentation and classification of nuclei in multi-tissue histology images. *Med. Image Anal.* **58**, 101563 (2019).

3.	Chen, L. C., Zhu, Y., Papandreou, G., Schroff, F. & Adam, H. Encoder-decoder with atrous separable convolution for semantic image segmentation. in *Lecture Notes in Computer Science (including subseries Lecture Notes in Artificial Intelligence and Lecture Notes in Bioinformatics)* vol. 11211 LNCS 833–851 (2018).

4.	Baheti, B., Innani, S., Gajre, S. & Talbar, S. Eff-UNet: A novel architecture for semantic segmentation in unstructured environment. *IEEE Comput. Soc. Conf. Comput. Vis. Pattern Recognit. Work.* **2020**-**June**, 1473–1481 (2020).

5.	Cao, H. *et al.* Swin-Unet: Unet-Like Pure Transformer for Medical Image Segmentation. *Lect. Notes Comput. Sci. (including Subser. Lect. Notes Artif. Intell. Lect. Notes Bioinformatics)* **13803 LNCS**, 205–218 (2023).

6.	Bilal, M. *et al.* Development and validation of a weakly supervised deep learning framework to predict the status of molecular pathways and key mutations in colorectal cancer from routine histology images: a retrospective study. *Lancet Digit. Heal.* **3**, e763–e772 (2021).

7.	Shephard, A. J. *et al.* A fully automated and explainable algorithm for predicting malignant transformation in oral epithelial dysplasia. *npj Precis. Oncol.* **8**, (2024).

8.	Macenko, M. *et al.* A method for normalizing histology slides for quantitative analysis. *Proc. - 2009 IEEE Int. Symp. Biomed. Imaging From Nano to Macro, ISBI 2009* 1107–1110 (2009) doi:10.1109/ISBI.2009.5193250.

9.	Ganin, Y. *et al.* Domain-adversarial training of neural networks. in *Advances in Computer Vision and Pattern Recognition* vol. 17 189–209 (2017).

10.	He, K., Zhang, X., Ren, S. & Sun, J. Identity mappings in deep residual networks. *Lect. Notes Comput. Sci. (including Subser. Lect. Notes Artif. Intell. Lect. Notes Bioinformatics)* **9908 LNCS**, 630–645 (2016).

11.	Shephard, A. J. *et al.* Simultaneous Nuclear Instance and Layer Segmentation in Oral Epithelial Dysplasia. *Proc. IEEE/CVF Int. Conf. Comput. Vis. Work.* **October**, 552–561 (2021).

12.	Pocock, J. *et al.* TIAToolbox as an end-to-end library for advanced tissue image analytics. *Commun. Med.* **2**, 120 (2022).

13.	Aubreville, M. *et al.* Mitosis domain generalization in histopathology images — The MIDOG challenge. *Med. Image Anal.* **84**, 102699 (2023).

14.	Jahanifar, M. *et al.* Stain-Robust Mitotic Figure Detection for the Mitosis Domain Generalization Challenge. *arXiv* 3–5 (2021).

15.	Jahanifar, M. *et al.* Mitosis Detection, Fast and Slow: Robust and Efficient Detection of Mitotic Figures. *Med. Image Anal.* **94**, 103132 (2024).

16.	Bashir, R. M. S. *et al.* A digital score of peri-epithelial lymphocytic activity predicts malignant transformation in oral epithelial dysplasia. *J. Pathol.* (2023) doi:10.1002/path.6094.

17.	Campanella, G. *et al.* Clinical-grade computational pathology using weakly supervised deep learning on whole slide images. *Nat. Med.* **25**, 1301–1309 (2019).

18.	Ilse, M., Tomczak, J. M. & Welling, M. Attention-based deep multiple instance learning. *35th Int. Conf. Mach. Learn. ICML 2018* **5**, 3376–3391 (2018).

19.	Lu, M. Y. *et al.* Data-efficient and weakly supervised computational pathology on whole-slide images. *Nat. Biomed. Eng.* **5**, 555–570 (2021).
